# UDCA and INT-777 suppress cardiac fibrosis triggered by IL-11 through involvement of TGR5

**DOI:** 10.1101/2022.05.11.22274945

**Authors:** B. Reilly-O’Donnell, E. Ferraro, R. Tikhomirov, R. Nunez-Toldra, A. Shchendrygina, L. Patel, Y. Wu, A. L. Mitchell, A. Endo, L. Adorini, R. A. Chowdhury, P. K. Srivastava, F. S. Ng, C. M. Terracciano, C. Williamson, J. Gorelik

**Affiliations:** National Heart and Lung Institute, Imperial College London, London, United Kingdom; Department of Women and Children’s Health, King’s College London, London, United Kingdom; Intercept Pharmaceuticals Inc., New York, USA

## Abstract

Cardiac fibrosis occurs in a wide range of cardiac diseases and is characterised by the transdifferentiation of cardiac fibroblasts (FB) into myofibroblasts (MFB). Myofibroblasts produce large quantities of extracellular matrix proteins, resulting in myocardial scar. The antifibrotic effect of the bile acid ursodeoxycholic acid (UDCA) is established in cases of liver fibrosis but not the adult myocardium.

Our hypothesis is: UDCA is antifibrotic in the adult heart, mediated by the membrane bile acid receptor Takeda G protein-coupled receptor 5 (TGR5).

We constructed a predictive network of fibrosis using RNA-seq datasets. We found that UDCA and it’s analogue INT-777, both reduced MFB markers in rat and human FBs and living myocardial slices (LMS). Utilising a knock-out mouse model, we show that the antifibrotic effect of UDCA is mediated by TGR5. Finally, we performed RNA-seq upon UDCA-treated human FB and integrated with our network of fibrosis, establishing the mechanism of TGR5 agonists.

## Introduction

Cardiac fibrosis is a defining feature of maladaptive cardiovascular remodelling which occurs in a variety of cardiovascular diseases (CVDs) and associated conditions such as; obesity, diabetes and kidney disease (Boer et al., 2019). The development of a fibrotic scar is detrimental to myocardial performance; reducing contractility and altering electrical conduction (Venero et al., 2015; Cojan-Minzat et al., 2020), which can lead to heart failure (HF). To date, there are no effective anti-fibrotic treatments which are capable of regulating the fibrotic response. A treatment which targets cardiac fibrosis is therefore an urgent requirement for both prevention and treatment of heart failure.

Besides the reparative fibrosis occurring after direct myocardial injury, the importance of reactive or interstitial fibrosis in HF pathogenesis has been recognized (Sabbah et al., 1995). Evolving evidence suggests the contribution of endothelial dysfunction (Huby et al., 2015), chronic low-grade systemic, and myocardial inflammation to the development of interstitial cardiac fibrosis (Frantz et al., 2018). In the setting of chronic myocardial inflammation, activated pro-inflammatory resident macrophages (Frantz et al., 2018) support the release of pro-inflammatory cytokines and chemokines including transforming growth factor-beta (TGF-β1), a key regulator of the fibrotic process (Yousefi et al., 2020). Among the multifactorial pro-inflammatory actions of TGF-β1, it facilitates fibroblast (FBs) activation, resulting in their transdifferentiation into myofibroblasts, particularly through IL-11 signalling (Schafer et al., 2017; Corden et al., 2020). The appearance of MFBs is a crucial way-marker in the development of maladaptive myocardial fibrosis (Tarbit et al., 2019). Once they appear, MFBs persist, contributing to the dysregulation of extracellular matrix (ECM). MFBs are incredibly active, causing disturbance in the synthesis and degradation of cytokines, growth factors, and matrix metalloproteinases (MMPs) (Travers et al., 2016). This leads to excessive ECM component secretion, which facilitates interstitial fibrosis formation. The conversion of fibroblasts into myofibroblasts therefore represents an attractive target for anti-fibrotic treatments.

Ursodeoxycholic acid (UDCA) (3,7-dihydroxy-5-cholanic acid), a hydrophilic secondary bile acid (BA), is commonly used to treat primary biliary cholangitis (Lindor et al., 1994; Cheung et al., 2016) and intrahepatic cholestasis of pregnancy (Carey and Lindor, 2012; Farooqui et al., 2022). UDCA has been identified as an agonist of the farnesoid X receptor (Mueller et al., 2015), TGR5 (Ibrahim et al., 2018) and free fatty acid 4 receptors (Xu et al., 2022). Besides its hepatoprotective, anti-apoptotic(Rodrigues and Steer, 2001; Amaral et al., 2009) and antioxidant effects (Lapenna et al., 2002), recent findings have demonstrated the anti-inflammatory (Tanaka et al., 2015; Calmus and Poupon, 1991) and antifibrotic potential of UDCA in the liver. The effect of BAs upon the heart was first described by Williamson et al. in 2001, since this initial publication the mechanism of action of BAs upon myocardial tissue has been extensively investigated (Williamson et al., 2001). UDCA has been identified to have cardioprotective effects against taurocholic acid-induced arrhythmia (Gorelik et al., 2004) and to modulate the action potential of cardiomyocytes and myofibroblasts (Schultz et al., 2016; Miragoli et al., 2011). Several studies have proposed that UDCA and its conjugate tauroUDCA (TUDCA) may have both anti-inflammatory and anti-fibrotic effects in the heart (von Haehling et al., 2012; Rani et al., 2017). In a mouse model of left ventricle pressure overload, induced by transverse aortic constriction (TAC), oral administration of TUDCA significantly reduced collagen deposition in the myocardium (Rani et al., 2017). Interestingly, the levels of pro-inflammatory proteins (TGF-β and p-Smad3) and mRNA expression of ECM proteins (Collagen 1α1 and 3α1) were decreased in the myocardial tissue of TUDCA-treated mice (Rani et al., 2017). The ability of UDCA to reduce plasma levels of pro-inflammatory cytokines was also confirmed in a small clinical study (von Haehling et al., 2012). In HF patients, UDCA treatment was associated with a lower concentration of soluble tumour necrosis factor α-receptor 1 (TNFα -1) in plasma, whereas the concentrations of TNFα and interleukin-6 remained unchanged (von Haehling et al., 2012). In 2016, for the first time, Schulz et al reported a significant decrease of MFBs in neonatal rat and human FBs, cultured in hypoxic conditions after UDCA treatment (Schultz et al., 2016). Recently our group has identified that UDCA, along with many other bile acids, can cause an increase in intracellular cyclic-AMP in neonatal rat ventricular myocytes (Ibrahim et al., 2018), this effect was attributed to the activation of the receptor GPBAR1/ TGR5. However, the anti-fibrotic action of UDCA in adult human culture models of cardiac fibrosis, requires further investigation. Additionally, the mechanism of anti-fibrotic action of UDCA in the heart requires further exploration.

TGR5 is a G-coupled protein coupled membrane receptor which can be activated by BAs, it is currently being investigated as a potential target for anti-fibrotic treatments (Pols et al., 2011). Mapping studies indeed indicate that the TGR5 receptor is broadly expressed in human and animal tissue and organs, including the heart (Kawamata et al., 2003). This is not limited to cardiomyocytes, TGR5 has been found in endothelial (Keitel et al., 2007) and immune cells (Perino et al., 2014)– key regulators of the fibrotic process. However, little is known about TGR5’s expression in cardiac fibroblasts. Bile acid activation of TGR5 induces cyclic adenosine monophosphate (cAMP) production, stimulation of various intracellular signalling into the cytoplasm such as AKT (Kida et al., 2013; Perino et al., 2014), extracellular signal-regulated kinases 1 / 2 (Masyuk et al., 2013a), and NF-κB pathways (Pols et al., 2011; Wang et al., 2011; Yoneno et al., 2013). This leads to downstream signalling events that contribute to the regulation of basal metabolism, inflammatory response, and tissue regeneration. Current knowledge suggests that activation of the TGR5 receptor by the selective agonist INT-777 (Sato et al., 2008) reduces the expression of pro-inflammatory cytokines including TGFβ-1 by glomerulus mesangial cells in kidney (Yang et al., 2016) and decreases renal fibrosis in diabetic mice (Wang et al., 2016). However, the physiological role of TGR5 receptors in the heart still requires investigation.

In this study, we investigated the anti-fibrotic effect of UDCA in human and rat cell culture models of cardiac fibrosis. We also established the mechanism of action of UDCA in cardiac FBs isolated from TGR5 KO mice. We utilised human and rat myocardial slices to translate our culture studies into a multi-cellular model. Finally, we used published RNAseq datasets of healthy and DCM patients to confirm our mechanism of action of TGR5 agonists as a treatment of cardiac fibrosis.

## Results

### Prediction of interaction of GPBAR1 with profibrotic network in human DCM FBs

A network of genes differentially expressed due to IL-11 or TGF-β treatment of human FBs was constructed from the RNA-seq datasets published by Schafer et al. (Schafer et al., 2017) and Chen et al. (Chen et al., 2019). Analysis of the gene ontology enrichment of these datasets showed a high prevalence of genes clearly associated with cardiac fibrosis, in the IL-11 (Fig. 1A) and WWP2/ TGF-β networks (Fig. 1B). The ‘profibrotic’ gene network was combined with known protein-protein interactions of GPBAR1 (the gene encoding TGR5) (Fig. 1C) to construct a predictive network of the interaction of GPBAR1 / TGR5 with profibrotic pathways. This provides a comprehensive prediction of how TGR5 interacts with pro-fibrotic networks.

**Fig. 1:**
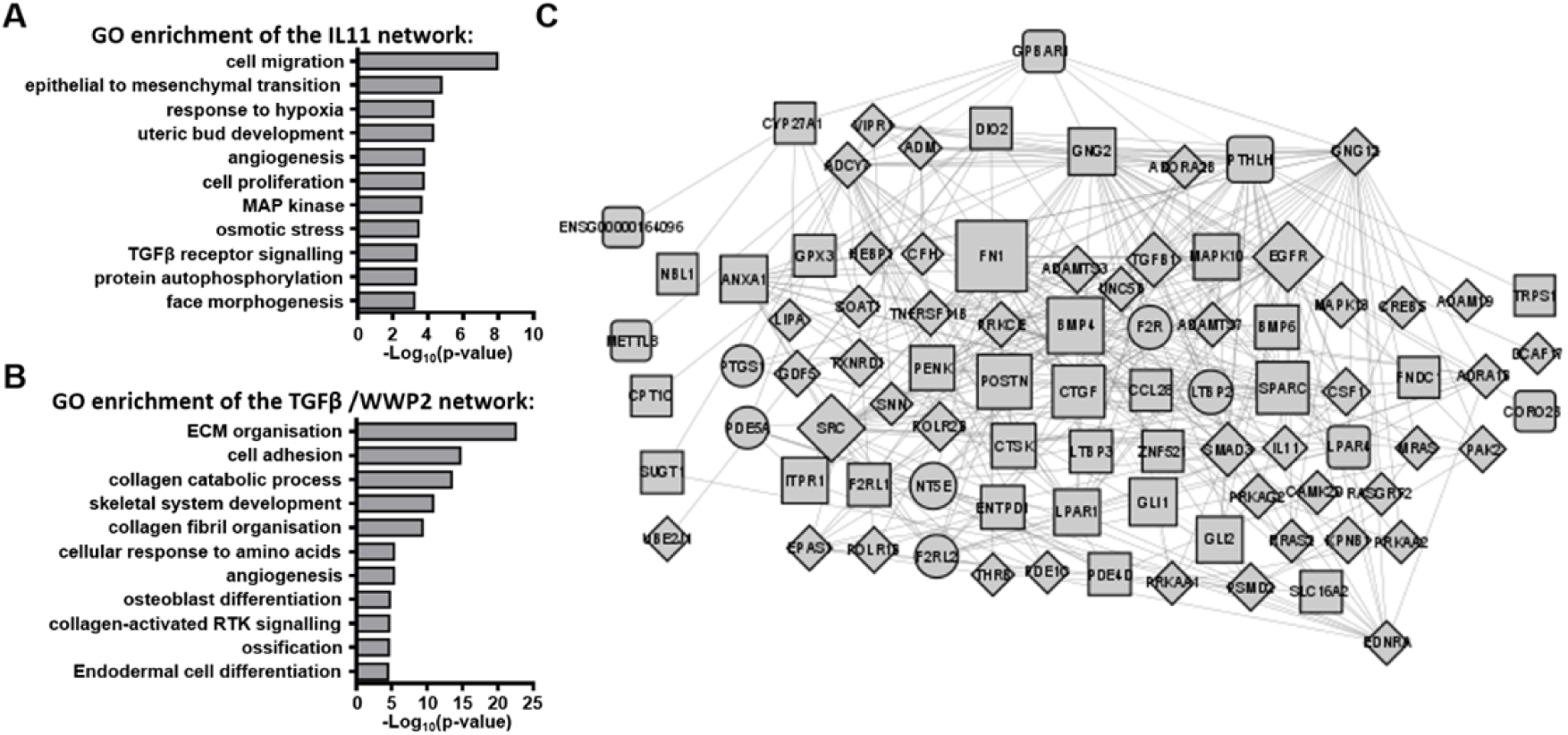
Construction of a fibrosis network. **A** Gene ontology enrichment of IL-11 network. **B** Gene ontology (GO) enrichment of TGF-β / WWP2 network. **C** A gene expression network constructed from published RNA-seq datasets and known interactors of GPBAR1. Circles represent genes belonging to the IL-11 co-expression network, diamonds represent genes belonging to the WWP2/ TGF-β network, squares represent genes belonging to both pro-fibrotic networks. Two levels of the network are presented.

### RNA-seq analysis reveals antifibrotic pathways associated with TGR5 signalling

We performed RNA-seq analysis upon cultured human DCM FBs. Incubation of cells with IL-11 caused a significant change (p-value< 0.05 or -log10(p-value)>1.42) in the expression in 360 genes (Fig. 7A). Pre-treatment of cultures with 1μM UDCA caused a reversal in the expression patterns (Fig. 7B). A plot of log10-fold changes of 109 genes which were significantly regulated in both IL-11-treated and UDCA-treated conditions (Fig. 7C). The Log(fold change) of genes in response to UDCA was mapped (Fig. 7D) onto the predictive network (established in Fig. 1A). Gene set enrichment analysis showed a reversal of the IL-11 (Fig. 7E) and WWP2/ TGF-β (Fig. 7F) profibrotic pathways (false discovery rate (adjusted p-value< 0.05)).

### UDCA prevents the expression of cardiac fibrosis markers in wild type (WT) rat and human DCM FBs

Rat ventricular FBs pre-treated with UDCA for 24hrs, before stimulation by 5ng/ml IL-11, showed a concentration-dependent decrease in the percentage of α-smooth muscle actin (αSMA) positive cells (Fig. 2A). UDCA significantly reduced the percentage of αSMA positive cells from 57.7 ±2.2% to 35.8 ±4.0% when pre-treated with 10μM UDCA (Fig. 2B). The percentage of αSMA positive cells when pre-treated with 1μM UDCA was also significantly reduced from IL-11 treated controls (Fig. 2B).

**Fig. 2.**
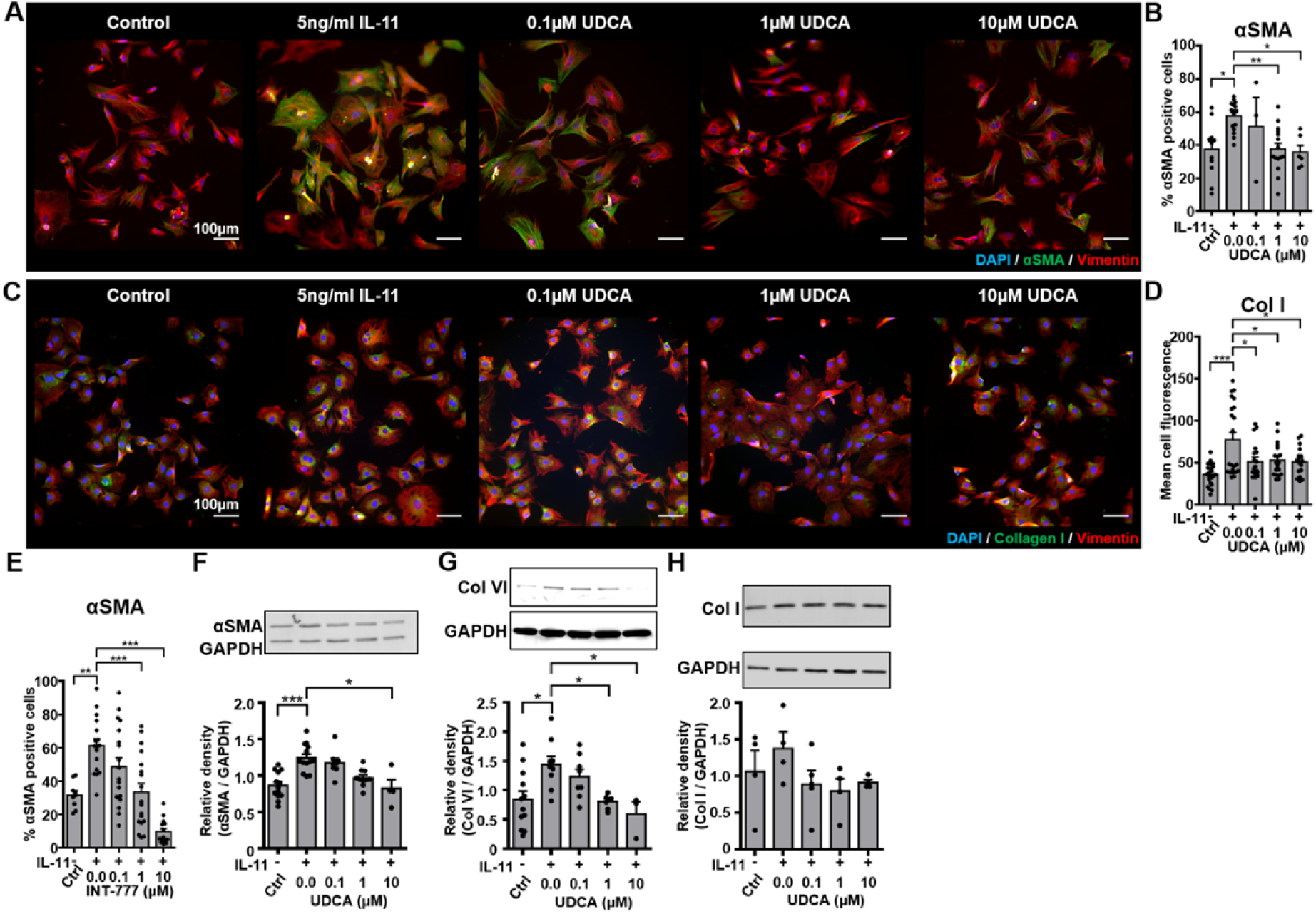
UDCA is antifibrotic in WT adult rat fibroblasts. **A** Representative images of IL-11-treated WT rat FBs stained for αSMA (M0851, Dako) green, Vimentin (PA1-16759, Thermo) red and DAPI blue. Culture conditions are displayed above each image. **B** Percentage of cells positive for αSMA staining in response to UDCA. n= 3-16. **C** Representative images of FBs stained for Collagen I (ab34710, abcam) green, Vimentin (PA1-16759, Thermo) red and DAPI blue. Culture conditions are above each image. **D** Mean cell fluorescence of Collagen I staining in response to UDCA. n= 17-22. **E** Percentage of cells positive for αSMA staining in response to INT-777. n= 9-19. **F** Representative Western blot and quantification of WT rat cell lysate probed for αSMA and GAPDH (2118, Cell Signalling Technologies) in response to UDCA. n=4-13. **G** Representative Western blot and quantification of WT rat cell lysate probed for Collagen VI (ab6588, abcam) and GAPDH in response to UDCA. n=3-9. **H** Representative Western blot and quantification of WT rat lysate probed for Collagen I (ab34710) and GAPDH in response to UDCA. n= 4-5.

Similarly, collagen I staining of WT rat FBs was significantly reduced by pre-treatment of cultures with UDCA (Fig. 2C). Mean cell fluorescence was reduced from 77.3 ±9.0 to 51.6 ±4.1 when cells were incubated with 10μM UDCA (Fig. 2D). The mean cell fluorescence of cells pre-treated with 0.1 and 1μM UDCA was also significantly reduced from IL-11 treated controls; there was no significance between UDCA concentrations tested.

Pre-treatment of WT rat FBs with INT-777 significantly reduced the percentage of αSMA positive cells in a concentration-dependent manner. Incubation of FBs with 10μM INT-777 significantly reduced the percentage of αSMA positive cells from 61.5 ±4.0% to 9.7 ±2.0%, pre-incubation of cells with 1μM INT-777 also significantly reduced the percentage of αSMA positive cells compared to IL-11 treated control (Fig. 2E).

Western blot analysis of protein expression in WT rat FBs identified significant reduction in αSMA (Fig. 2F) and Collagen VI (Fig. 2G) at both 1μM and 10μM. There was no significant change in Collagen I expression observed by Western blot analysis (Fig. 2H).

UDCA reduced the percentage of αSMA positive human DCM FBs in a concentration-dependent manner (Fig. 3A). Pre-incubation of cells with either 1μM or 10μM UDCA significantly reduced αSMA positive cells from 44.0 ±3.0% to 21.7 ±2.8% and 16.7 ±2.6% respectively (Fig. 3B). However, we did not observe any significant change in collagen I staining (Fig. 3C and D).

**Fig. 3.**
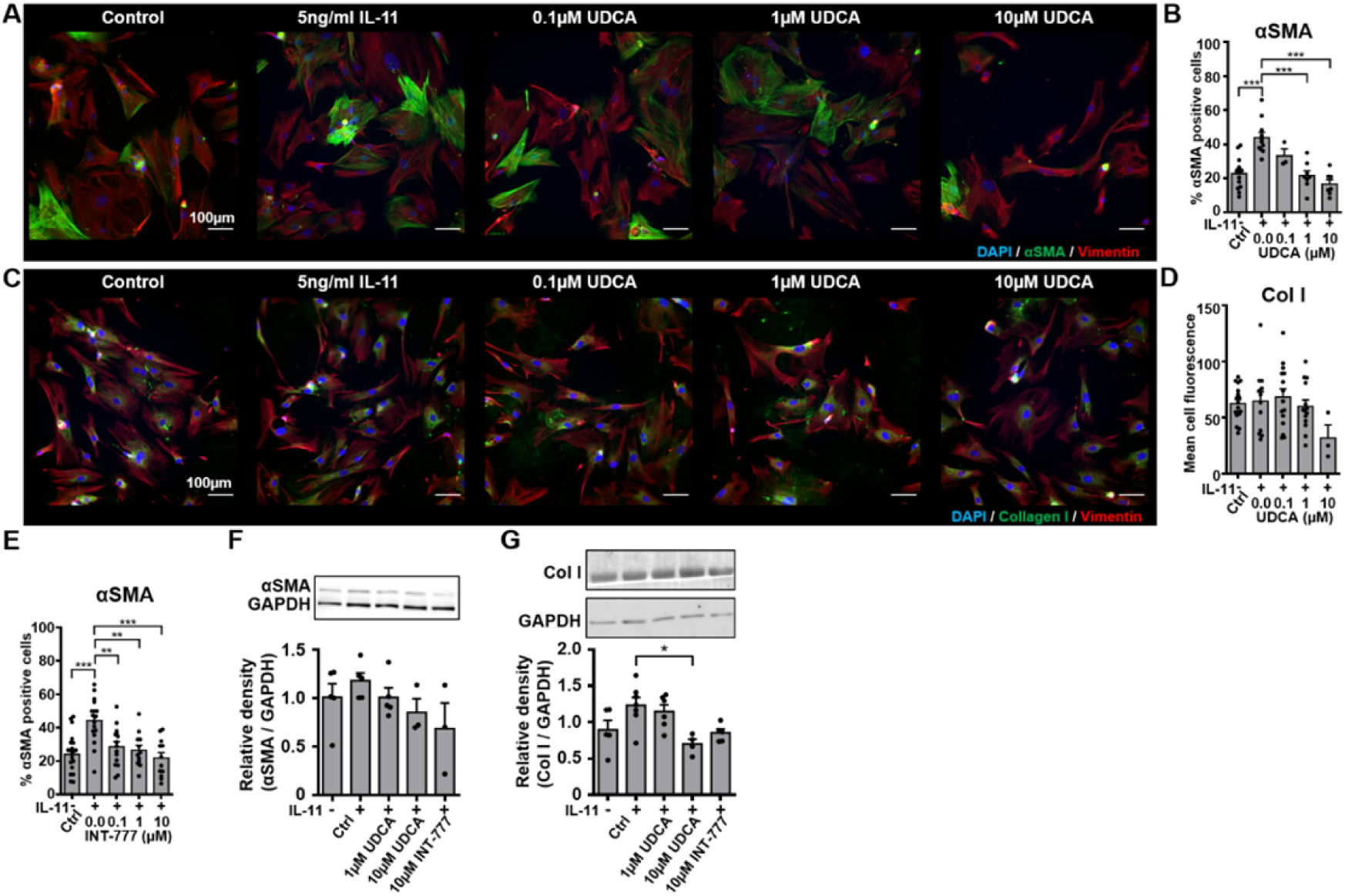
UDCA is antifibrotic in human dilated cardiomyopathy cardiac fibroblasts. **A** Representative images of human DCM FB stained for αSMA (M0851, Dako) green, Vimentin (PA1-16759, Thermo) red and DAPI blue. Culture conditions are displayed above each image. **B** Percentage of cells positive for αSMA staining in response to UDCA. n= 3-13. **C** Representative images of FBs stained for Collagen I (ab34710, abcam) green, Vimentin (PA1-16759, Thermo) red and DAPI blue. Culture conditions are above each image. **D** Mean cell fluorescence of Collagen I staining in response to UDCA. n= 3-18. **E** Percentage of cells positive for αSMA staining in response to INT-777. n= 12-21. **F** Representative Western blot and quantification of human DCM cell lysate probed for αSMA and GAPDH (2118, Cell Signalling Technologies) in response to UDCA or INT-777. n= 3-5. **G** Representative Western blot and quantification of human DCM cell lysate probed for Collagen I (ab34710) and GAPDH in response to UDCA or INT-777. n=4-7.

A concentration-dependent decrease of αSMA positive human DCM FBs was also observed in response to incubation with INT-777 (Fig. 3E). A significant reduction in αSMA positive cells was observed when FBs were incubated with 10μM INT-777 from 44.3 ±3.19% to 21.9 ±3.3%, both 0.1μM and 1.0μM INT-777 also significantly reduced the percentage of αSMA positive cells when compared to IL-11 treated control (Fig. 3E).

There was a trend, but no significant reduction, in αSMA expression of human DCM FBs pre-treated with UDCA or INT-777 at any concentration tested, when assessed by Western blot analysis (Fig. 3F). However, Collagen I expression was significantly reduced following treatment with 10μM UDCA (Fig. 3G).

### UDCA reduces fibrosis markers and improves contractility of WT rat living myocardial slices

Rat living myocardial slices were produced and cultured with electrical stimulation for 48hrs using established methods (Perbellini et al., 2018; Watson et al., 2017; Pitoulis et al., 2020) and then stained for Collagen I (Fig. 4A) and assessed by percentage area stained (Fig. 4B). Incubation of rat slices with 10ng/ml IL-11 significantly increased the percentage area of Collagen I compared to untreated control slices from 12.4 ±0.9% to 16.0 ±0.9. Co-treatment of slices with 10μM UDCA and IL-11 significantly reduced the percentage area of Collagen I from 16.0 ±0.9 to 9.7 ±0.9%. Incubation of slices with UDCA alone had no significant effect upon the area of collagen I staining, although there is a trend to a reduced area of staining (9.3 ±0.5%). The reduction of collagen I staining was also reflected by Western blot (Fig. 4C), where co-treatment of slices with 10μM UDCA and IL-11 reduced the expression of collagen I. The functionality of live rat LMS was assessed by a force-transducer with pacing at 1Hz (Fig. 4D). Maximal contractility of slices was reduced by IL-11 from 3.8 ±0. mN/mm^2^ to 1.3 ±0.2 mN/mm^2^. Co-incubation of the slice with 10μM UDCA and IL-11 improved maximal contractility from 1.3 ±0.2 mN/mm^2^ to 3.8 ±0.3 mN/mm^2^ (Fig. 4E). Co-treatment of slices with 10μM UDCA and IL-11 reduced the half-width of contractions from 244.5 ±7.4ms to 160.6 ±9.9ms (Fig. 4F).

**Fig. 4.**
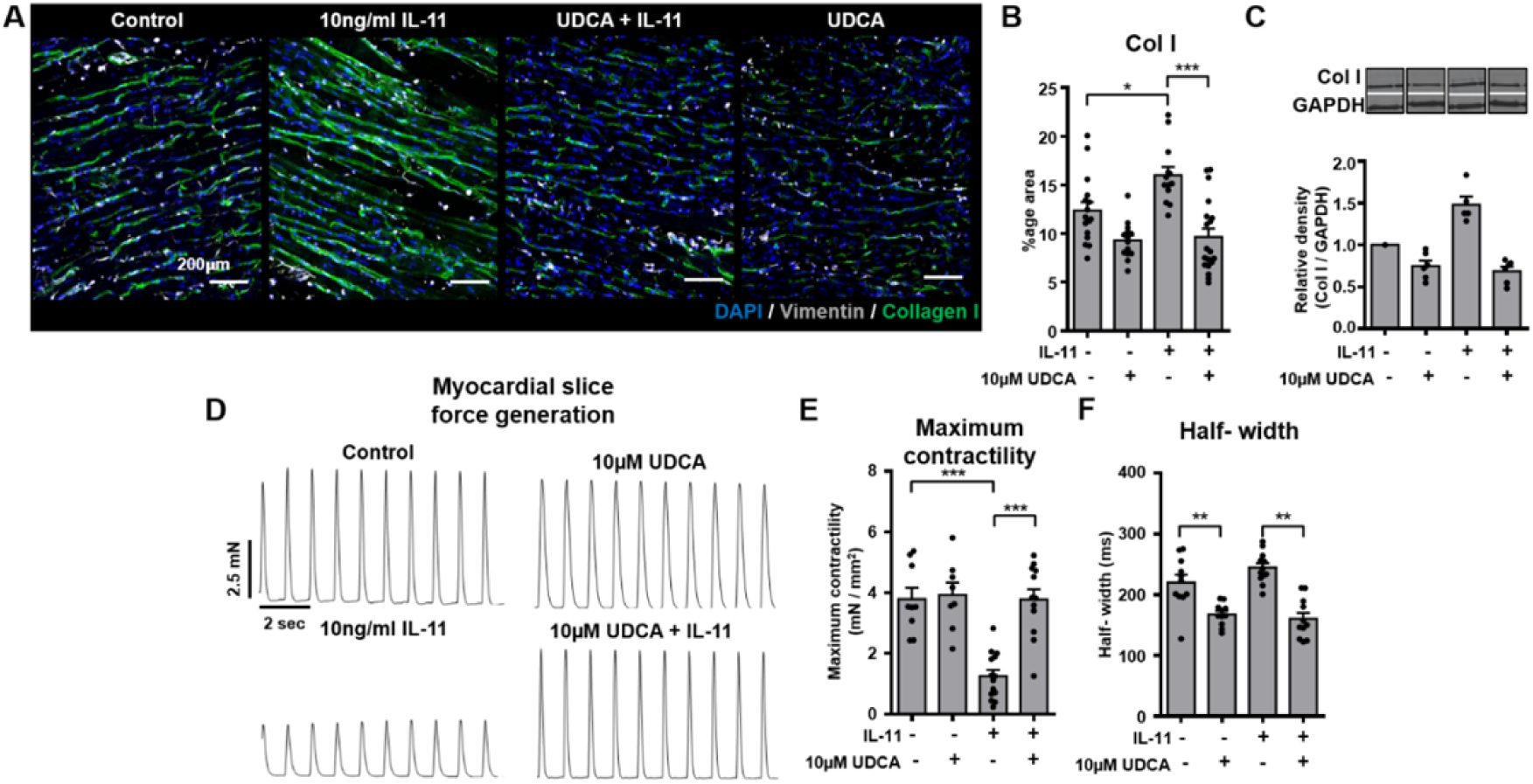
UDCA reduces markers of fibrosis and improves contractile function of adult rat living myocardial slices. **A** Representative images of IL-11-treated WT rat LMS stained for Collagen I (ab34710, abcam) green, Vimentin (PA1-16759, Thermo) grey and DAPI blue. Culture conditions are displayed above each image. **B** Percentage area of collagen I staining of LMS. n= 13-19, N= 3-7. **C** Representative Western blot and quantification of LMS lysate probed from Collagen I. n=5-6. **D** Representative contractile activity of WT rat LMS. Culture conditions are displayed above each trace. **E** Maximum contractility of LMS. n=8-14, N=6-8. **F** Contractility half-width. n= 11-12, N=6-8.

### UDCA reduces markers of fibrosis in human DCM living myocardial slices

The percentage area of collagen I staining in human LMS was significantly higher in human DCM slices compared to donor (18.5 ±1.1% vs. 11.3 ±1.4%) (Fig. 5A and B). “Donor” slices were produced from organ donor non-failing hearts not suitable for transplantation. Treatment of human DCM LMS with 10μM UDCA for 48hrs significantly reduced the percentage collagen area from 18.5 ±1.1% to 13.5 ±0.8% (Fig. 5A and B). The contractility of slices was also assessed (Fig. 5C), LMS prepared from DCM hearts had very low contractility (9.7 ±2.7% of donor LMS contractility). The relative contractility of human DCM slices was slightly increased when slices were treated with 10μM UDCA for 48hrs (12.9 ±3.9% vs. 9.7 ±2.7% of donor LMS contractility), however this was not significant (Fig. 5D).

**Fig. 5.**
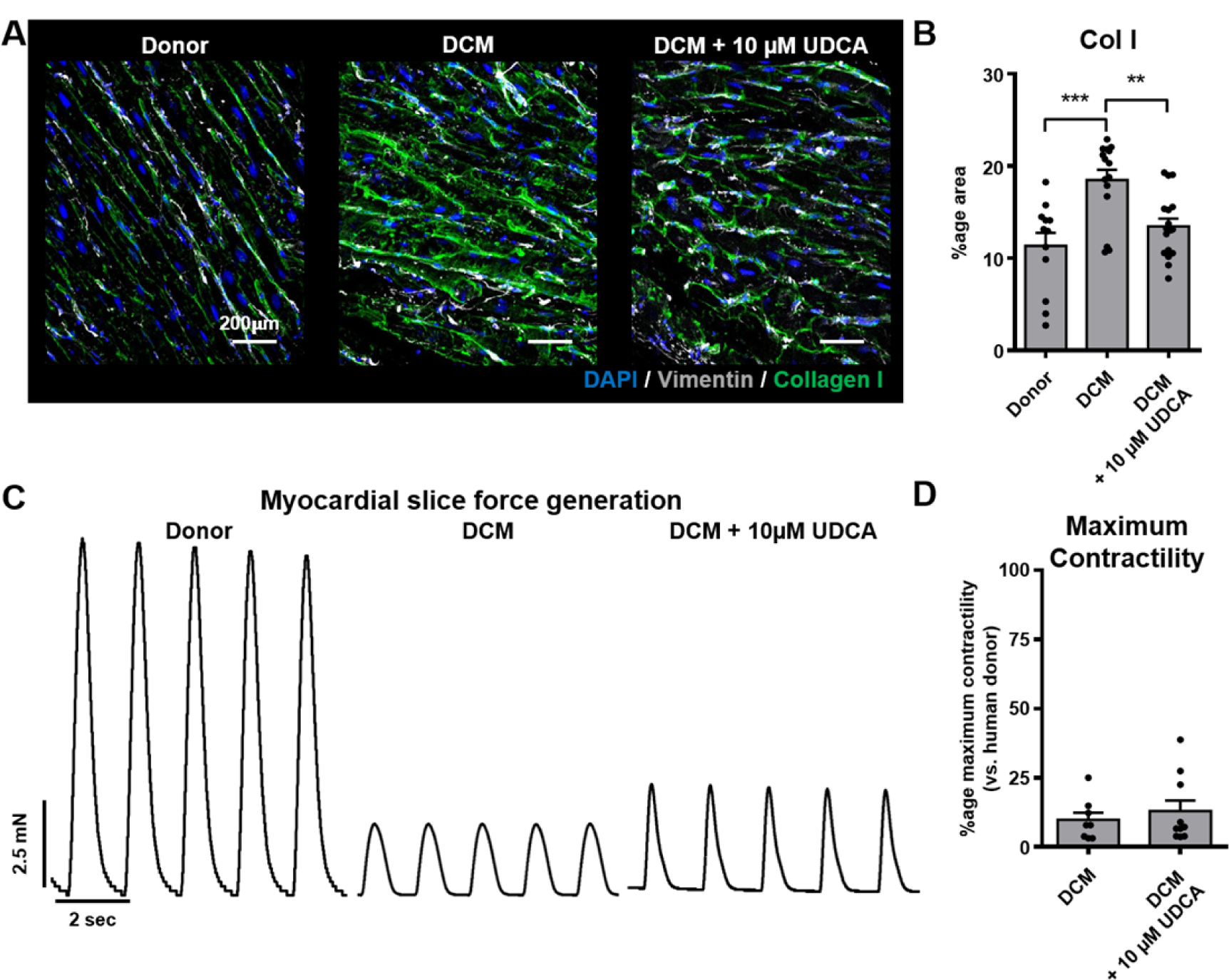
UDCA reduces markers of fibrosis in human living myocardial slices. **A** Representative images of human LMS stained for Collagen I (ab34710, abcam) green, Vimentin (PA1-16759, Thermo) grey and DAPI blue. Culture conditions are displayed above each image. **B** Percentage area of collagen I staining of LMS. n= 12-17, N=3-7. **C** Representative contractile activity of LMS. Culture conditions are displayed above each trace. **D** Maximum contractility of human LMS normalised to average human donor contractility. n=8-10, N=3-7.

### Knock-out of TGR5 reduces the anti-fibrotic effect of UDCA in mouse fibroblasts

Incubation of WT mouse FBs with 1μM and 10μM UDCA significantly reduced the percentage of αSMA positive cells from 56.6 ±8.5% to 30.4 ±7.9% and 22.9 ±2.3% respectively (Fig. 6A and B). There was no significant reduction of αSMA positive cells in TGR5 KO FBs, when pre-treated with 1 or 10μM UDCA (Fig. 6B). Mean cell fluorescence of both WT and TGR5 KO FBs stained for collagen I was reduced in both cell types when incubated with 10μM UDCA (Fig. 6C). The TGR5-specific agonist, INT-777, also caused a reduction in the percentage of αSMA expressing WT FBs. This effect of INT-777 was lost in TGR5 KO FBs (Fig. 6D). Western blot analysis of αSMA expression reflected the findings of our imaging experiments (Fig. 6E). There was no change in total ERK1/2 expression in any conditions examined (Fig. 6F). Analysis of ERK1/2 phosphorylation in WT mouse FBs identified that pre-treatment of WT FBs with 1μM UDCA significantly reduced phosphorylation (relative density was reduced from 1.3 ±0.1 to 1.0 ±0.1) (Fig. 6G). This reduction in ERK1/2 phosphorylation by UDCA was lost in TGR5 KO FBs (Fig. 6G).

**Fig. 6.**
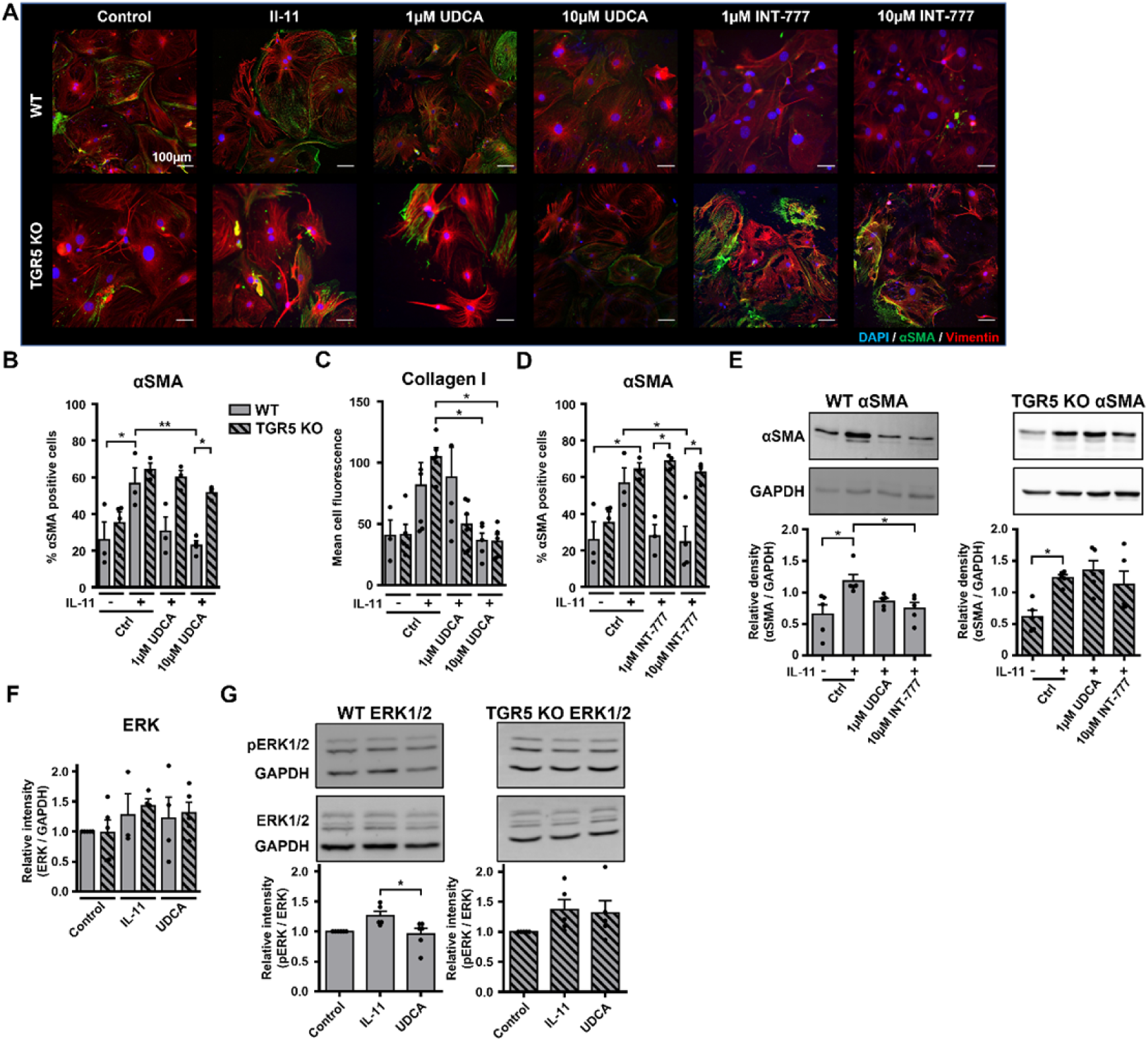
Reduction of fibrosis markers by UDCA is abolished in TGR5 KO mouse fibroblasts. **A** Representative images of mouse FBs stained for αSMA (M0851, Dako) green, Vimentin (PA1-16759, Thermo) red and DAPI blue. Culture conditions are displayed above each image. **B** Percentage of αSMA positive FB in response to UDCA. n= 3-4. **C** Mean cell fluorescence of collagen I (ab34710, abcam). n=4-6. **D** Percentage of αSMA positive FB in response to INT-777. n= 3-4. **E** Representative Western blot and quantification of mouse cell lysate probed for αSMA and GAPDH (2118, Cell Signalling Technologies) in response to UDCA or INT-777. n=4-5. **F** Quantification of western blot for total ERK1/2 (4695, Cell Signalling Technologies) n= 3-5. **G** Representative Western blot and quantification of mouse cell lysate probed for ERK1/2 (4695, Cell Signalling Technologies), phosphorylated ERK1/2 (4370, Cell Signalling Technologies) and GAPDH in response to UDCA. n=5-6.

## Discussion

Using multiple models of cardiac fibrosis, we have demonstrated the antifibrotic effect of the TGR5 agonists UDCA and INT-777. We have characterised the interaction of the pro-fibrotic network with TGR5 and suggest that UDCA and other TGR5 agonists have the potential to reduce cardiac fibrosis.

### Antifibrotic effect of bile acids in adult rat models of cardiac fibrosis

The antifibrotic effect of UDCA and INT-777 in cultured adult rat FB was identified by immunostaining and Western blot. UDCA induced a significant reduction in α-SMA staining (Fig. 2B) which was reflected in Western blots (Fig. 2F). This was not the case for Collagen I, where staining was significantly reduced by UDCA (Fig. 2C and D) but not paralleled in Western blot (Fig. 2H). Interestingly, we were able to detect a reduction in the anchoring collagen type VI by Western blot analysis when cells were pre-treated with UDCA (Fig. 2G). These data taken together indicate that pre-treatment of cultures with UDCA before stimulation with IL-11 prevents the activation of FB and therefore the emergence of MFB in cultures. We also found that the TGR5-specific agonist, INT-777 (Pellicciari et al., 2009), reduced the number of α-SMA positive cells (Fig. 2E) to an even greater extent than UDCA, indicating that it too can prevent the transdifferentiation of FB into MFB. This correlates with similar experiments investigating liver fibrosis, suggesting a common mechanism of action (Ye et al., 2020).

Transferring these cell culture experiments to multi-cellular and hetero-cellular LMS, we found that collagen I staining was reduced in slices treated with 10μM UDCA (Fig. 4A-C). This reduction of collagen I is indicative of a reduction of extracellular matrix particularly with interstitial fibrosis, rather than focal fibrosis. Assessment of the function of the LMS was determined by force transducer, there was a clear reduction in contractility of the slice when treated with IL-11 alone, but co-treatment with 10μM UDCA significantly increased function (Fig. 4E and F). The reduction in contractile dynamics of UDCA-only treated slices, compared to control, perhaps points towards some non-fibroblast mediated effects of UDCA (Fig. 4F). It is known that UDCA cannot influence contractility in neonatal mouse myocytes despite promoting cAMP release (Ibrahim et al., 2018) although there is clear evidence that UDCA can influence electrophysiological properties of the heart, and reduce ischaemia-induced arrythmias (Ferraro et al., 2020; Miragoli et al., 2011; Schultz et al., 2016; Gorelik et al., 2003). In our recent publication, this was attributed to increased phosphorylation of connexin-43 proteins (Ferraro et al., 2020), which perhaps explains the increased contraction dynamics of slices treated with UDCA.

### Antifibrotic effect of bile acids in human models of cardiac fibrosis

UDCA was found to inhibit the transdifferentiation of human FB into MFB (Fig. 3). Interestingly, unlike adult rat FB, there was a significant reduction in collagen I when assessed by Western blot but not immunostaining. There was a clear reduction in the number of MFBs in cultures treated with INT-777 (Fig. 3E), indicating that the antifibrotic action of the bile acids are mediated by TGR5.

Treatment of human failing (DCM) slices with UDCA significantly reduced the expression of collagen I (Fig. 5A and B), indicating that UDCA is antifibrotic in human slices as well as cultured FB. The reduction of collagen I in slices did not result in a significant increase in slice contractility (Fig. 5D). This is perhaps due to the samples available; cardiac tissue used to produce LMS is at an end stage of heart failure and so there is a reduction in the number of and function of myocytes in the slice. One may expect that reduced cardiac fibrosis will, however, result in improved diastolic function, countering the dysfunction caused by fibrosis (Moreo et al., 2009), further studies utilising LMS would be particularly useful in investigating this phenomenon. This indicates that any future treatments of heart failure involving bile acids would require administration early in the pathogenesis of heart failure if contractile function is to be maintained.

### TGR5 is required to prevent transdifferentiation of fibroblasts into myofibroblasts

The antifibrotic effect of UDCA was observed in WT mouse FB (Fig. 6) but was lost in TGR5 KO cultures. This agrees with our pharmacological study in which INT-777, the TGR5-specific agonist, prevented the expression of MFB markers (Fig. 2E, 3E). Our lab has previously identified that UDCA can stimulate cAMP release via TGR5 in neonatal rat myocytes (Ibrahim et al., 2018) and that unconjugated UDCA is more potent than tauro-or glyco-conjugated UDCA. Along with the present data we propose TGR5 activation as the mechanism of action of both UDCA and INT-777 in the inhibition of cardiac fibrosis. Interestingly, we found that UDCA significantly reduced the phosphorylation (but not expression) of ERK1/2 in WT, but not in TGR5 KO FB (Fig. 6F and G), identifying a downstream signalling of TGR5 in cardiac FB similar to ciliated cholangiocytes (Masyuk et al., 2013b).

### Role of TGR5 signalling in cardiac fibrosis

A gene expression network was constructed from published RNA-seq data (Schafer et al., 2017) and known TGR5 (GPBAR-1) interactors. In this network we identified genes regulated in pro-fibrotic conditions (stimulated by IL-11) and arranged them by their shortest path connectedness to TGR5/ GPBAR-1, three layers of the network are presented (Fig. 1C). Analysis of the previously published RNA-seq datasets using gene ontology annotations (DAVID) found a number of pathways expected to be activated in FB during proliferative fibrosis were indeed upregulated i.e. cell migration, cell proliferation, MAP kinase pathway, TGF-β signalling (Fig. 1A and B).

After performing our own RNA-seq experiments upon human DCM FB we found a number of genes to be significantly up-and down-regulated when cultures were stimulated with IL-11 (Fig. 7A). Interestingly pre-treatment of cultures with 1μM UDCA before stimulation with IL-11 caused an almost exact reversal of the gene expression profile (Fig. 7B), as noted by the scatter plot of significantly regulated genes in both conditions (Fig. 7C). Gene-set enrichment analysis, against the published IL-11 and WWP2/ TGF-β datasets showed that IL-11 enriched the profibrotic network and UDCA reversed this effect (Fig. 7D and E). Mapping our own experimental data onto our network (Fig. 7F) reveals the interaction of both the pro-fibrotic and anti-fibrotic pathways investigated in this study.

**Fig. 7.**
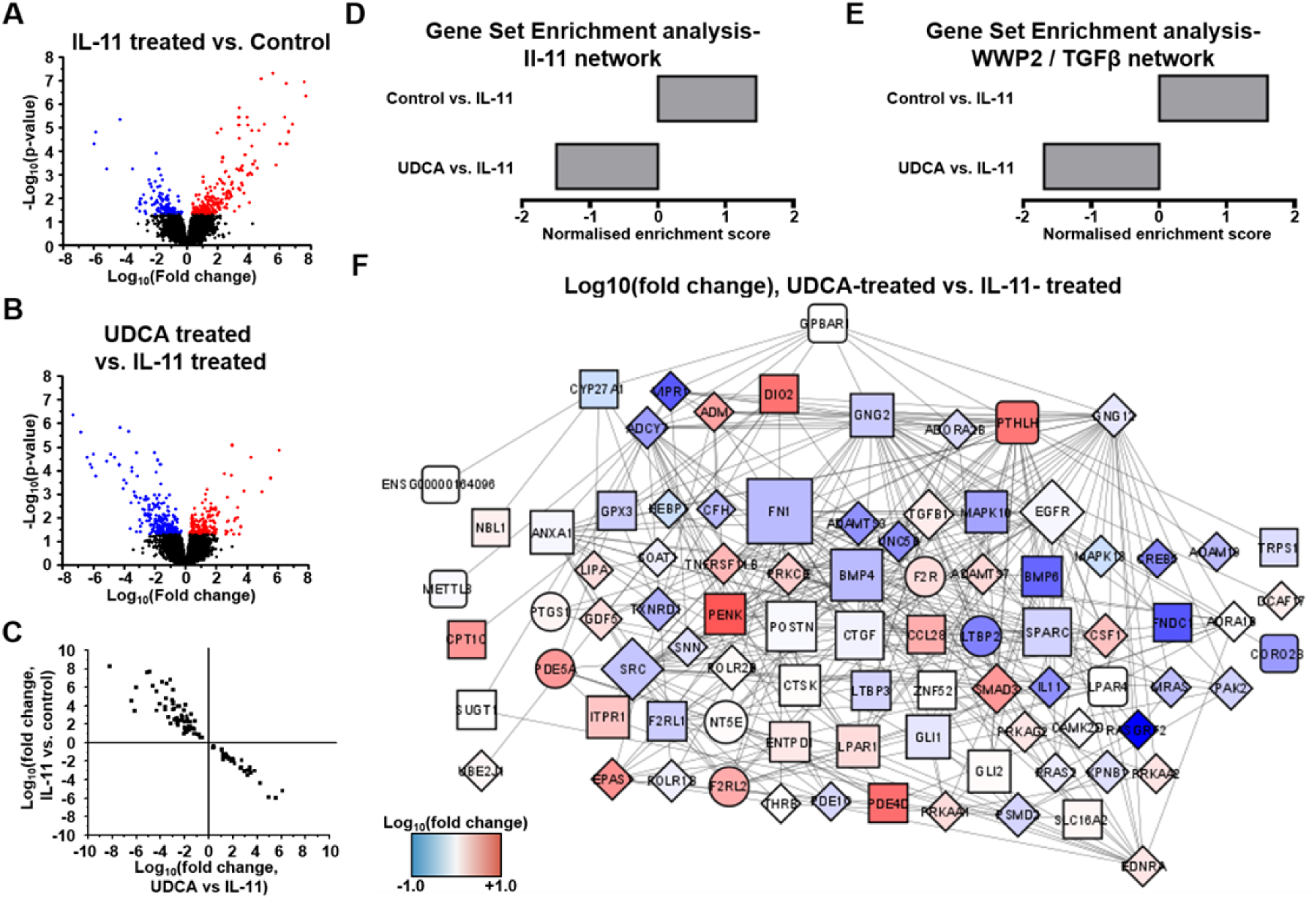
Identification of a cardiac fibrosis network in human fibroblasts. **A** Volcano plot of differentially expressed genes as determined by RNA-seq. Conditions compared were control (no treatment) vs 5ng/ml IL-11 stimulated human DCM FB. Significantly upregulated genes are marked in red, significantly down regulated genes are marked in blue. n=3. **B** Volcano plot of differentially expressed genes as determined by RNA-seq. Conditions compared were 5ng/ml IL-11 vs 1μM UDCA pre-treated stimulated human DCM FB. Significantly upregulated genes are marked in red, significantly down regulated genes are marked in blue. n=3. **C** Plot of significantly regulated genes of Human DCM FB. Log fold change in gene expression of IL-11 vs Control is plotted against UDCA vs IL-11. **D** Gene set enrichment analysis of IL-11 network. **E** Gene set enrichment analysis of WWP2 / TGF-β network. **F** Change in gene expression of fibrosis network generated in Fig. 1A due to treatment of human DCM cells with UDCA. Blue= reduced expression, Red= increased expression. Circles represent genes belonging to the IL-11 co-expression network, diamonds represent genes belonging to the WWP2/ TGF-β network, squares represent genes belonging to both pro-fibrotic networks. Two levels of the network are presented.

This study confirms the potential of UDCA and INT-777 as treatments of cardiac fibrosis in the adult myocardium. Using multiple animal and human models, we have identified that TGR5 agonists can prevent the transdifferentiation of FB into MFB and have characterised the gene expression network associated with the pro-and anti-fibrotic pathways.

### Ideas and speculation

Currently there are no drugs approved primarily for the treatment of cardiac fibrosis, despite a broad range of strategies investigated (Sweeney et al., 2020). Although inhibitors of the renin-angiotensin-aldosterone system have been shown to have beneficial effects on interstitial fibrosis, their effects are modest (Brilla et al., 2000). The repurposing of UDCA is safely prescribed in the treatment of primary biliary cholangitis (Parés et al., 2006) and intrahepatic cholestasis of pregnancy (Ovadia et al., 2021), would be of clinical value as an antifibrotic agent. There is already mounting evidence that TGR5 agonists are beneficial in patients with heart failure (von Haehling et al., 2012), ischaemia-induced arrythmia in rat (Ferraro et al., 2020) and TAC mice (Eblimit et al., 2018). What is unclear is how the non-fibroblast-mediated effects of TGR5 signalling may affect the long-term function of the heart and other organs. Unfortunately, TGR5 is widely expressed and so direct activation of the receptor may not represent a realistic drug target for cardiac fibrosis. The gene network produced as part of this study (Fig. 1C) may provide new targets which are more suited to a heart-specific treatment of cardiac fibrosis. Promisingly, there were no adverse events reported during two-week administration of UDCA in adult rats (Ferraro et al., 2020) or to patients for 4 weeks (von Haehling et al., 2012). Von Haehling et al. showed that administration of UDCA can increase peripheral blood flow (von Haehling et al., 2012), however there was no indication if this was due to improvement of cardiac function or peripheral vasculature. Another avenue would be to develop the TGR5-specific agonist INT-777 (over UDCA) as an anti-fibrotic agent, removing the potential ‘off-target’ effects of UDCA, for the treatment of cardiac fibrosis and perhaps administer this to patients with increased risk of developing fibrosis. Reducing cardiac fibrosis will likely have the benefits of improving diastolic function (Khalil et al., 2017) and also reducing the incidence of ventricular arrhythmias (Pahor et al., 1991), which accounts for a substantial proportion of deaths in patients with heart failure (Moss et al., 2002). To further understand the potential of TGR5 agonists as treatments of cardiac fibrosis there is a need to investigate these drugs in patients.

## Materials and methods

All reagents, unless stated, were purchased from Sigma-Aldrich (Dorset, UK).

INT-777 was provided by Intercept Pharmaceuticals, Inc. (New York, USA).

### Isolation and culture of rat and mouse cardiac fibroblasts

All animal experiments were performed in the United Kingdom (UK) according to the standards for the care and use of animal subjects determined by the UK Home Office (ASPA1986 Amendments Regulations 2012) incorporating the EU directive 2010/63/EU. The animal welfare and ethical review body committee of Imperial College London approved all protocols.

Rat (male, Sprague Dawley) and mouse (male, C57BL/6, WT and TGR5 KO) FBs were isolated by enzymatic digestion of ventricular tissue as previously described (Wright et al., 2018). FBs were found in the supernatant after centrifugal pelleting of cardiomyocytes and were cultured for no more than 20 days before cell fixation/ lysis.

Rat FBs were cultured in Dulbecco’s modified Eagle’s media (DMEM) supplemented with 10% fetal bovine serum (FBS) and 1% antibiotic-antimycotic solution, whereas mouse FBs were cultured in DMEM supplemented with 20% FBS and 1% antibiotic-antimycotic, at 37°C and 5% CO_2_.

Cultures were pre-treated for 24 hrs with UDCA or INT-777 before stimulation with 5ng/ml IL-11 (Rat; RPA057Ra01 (Caltag Med systems), Mouse; Z03052 (Genscript)) for a further 24 hrs.

### Isolation and culture of Human DCM fibroblasts

This study was supported by the supply of human failing hearts from the Cardiovascular Research Centre Biobank, at the Royal Brompton and Harefield hospitals (NRES Ethics number for biobank samples: 09/H0504/104+5, Biobank approval number: NP001-06-2015 and MED_CT_17_079) and donor hearts from NHS Blood and Transplant (NRES Ethics number 16/LO/1568, NHS Blood and Transplant study numbers 67 and 106). All procedures described were carried out in accordance with the Human Tissue Act 2004.

Left ventricular free-wall tissue was minced into small chunks (3mm x 3mm x 3mm) and then lightly digested in 0.05% Trypsin-EDTA for 2 min. Tissue chunks were then transferred into fibronectin-coated dishes and cultured in DMEM supplemented with 20% FBS and 1% antibiotic-antimycotic at 37°C and 5% CO_2_. When confluent, FB were harvested from the dishes and used for experiments.

Cultures were pre-treated for 24 hrs with UDCA or INT-777 before stimulation with 5ng/ml IL-11 (PHC0115, Life Technologies) for a further 24 hrs.

### Preparation and physiological monitoring of living myocardial slices

Living myocardial slices (LMS) were obtained from rat and human tissue, prepared and cultured in line with previous publications (Watson et al., 2017; Perbellini et al., 2018; Watson et al., 2019). Sprague-Dawley rats (300-350g) were anaesthetised by inhalation of 4% isoflurane at 4 L/min oxygen and then sacrificed by cervical dislocation. The heart was removed from the thoracic cavity and placed in ice-cold Tyrode’s slice solution (30 mM 2, 3-Butanedione Monoxime, 140 mM NaCl, 9 mM KCl, 10 mM Glucose, 10 mM HEPES, 1 mM MgCl_2_, 1 mM CaCl_2_, pH=7.4).

A 1.5 cm^2^ tissue block was dissected from the free wall of the left ventricle and, using a high precision vibratome (7000smz-2, Campden Instruments), 300μm thick LMS were prepared. The LMS were attached to custom-made 3D printed t-glase rectangular rings and then mounted on metallic stretchers which allow for the setting of diastolic load. In this study the LMS were cultured at physiological sarcomere length (SL)= 2.2 μm. LMS were placed inside sterile culture chambers and maintained in circulating, oxygenated media at 37°C for 48 hrs, under continuous electrical stimulation via carbon electrodes. Rat slices were paced at 1Hz (width 10ms, 15V) whereas human slices were paced at 0.5Hz (width 10ms, 15V). 10ng/ml recombinant rat IL-11 (Cloud-clone, USA) was used to induce the fibrotic phenotype, either in the absence or presence of 1μM UDCA.

LMS contractility was assessed using a force transducer (Harvard Apparatus, USA). After 48-hours in culture, LMS were connected to a force transducer, field stimulated, and progressively stretched in a stepwise manner, until the maximum isometric contraction was obtained. During contractility measurements LMS were perfused with oxygenated Tyrode’s solution at 37 °C. Recordings were obtained using AxoScope software and peak amplitude analysis conducted using Clampfit software (both Molecular Devices, USA).

### Western blot

Cell lysates were prepared in RIPA buffer supplemented with protease inhibitor cocktail and phosphatase inhibitor. Tissue lysates were snap frozen and then homogenised in SB-20 lysis buffer.

Samples were loaded on 8-10% polyacrylamide gels and ran at 100V for 1 hr. Proteins were transferred to PVDF membrane via semi dry transfer (Trans-Blot Turbo, Bio-Rad) or wet transfer at 100V for 1hr. Membranes were blocked with 5% (w/v) skimmed milk powder and then incubated with primary antibodies overnight (see figure legend). Secondary antibodies were either alexa-flour or HRP conjugated (see figure legend) and incubated with the membrane for 3 hrs. Blots were developed and/ or imaged using a Bio-Rad ChemiDoc MP.

### Immunostaining of isolated fibroblasts

Cells were cultured on glass coverslips. After the completion of experimental protocol, cells were fixed with 4% (v/v) PFA for 15 min on ice. Cells were permeabilised with 0.05% (v/v) Triton X-100 for 15 min and then blocked for 1 hr in 5% (w/v) BSA. Coverslips were incubated with primary antibody overnight, afterwards the coverslips were transferred into the appropriate secondary antibody-containing solutions for 1 hr. Coverslips were then mounted onto glass slides with ProLong Gold antifade mountant with DAPI.

Images were captured using either a Zeiss LSM-780 inverted confocal laser scanning microscope or Nikon Eclipse Ti with pE-4000 light source (Cool LED) and ORCA-Flash 4 camera (Hamamatsu). Imaging was assisted by the Facility for Imaging by Light Microscopy (FILM) at Imperial College London. FILM is part-supported by funding from the Wellcome Trust (grant 104931/Z/14/Z) and BBSRC (grant BB/L015129/1). Images were analysed with Fiji/ ImageJ.

### Immunostaining of living myocardial slices

LMS were washed in PBS and then fixed in 4% PFA. Slices were permeabilised with 1% Triton X-100 in blocking solution (10% fetal bovine serum, 5% bovine serum albumin and 10% horse serum in PBS) for 3 hrs at room temperature. Primary antibodies were diluted in PBS and incubated overnight at 4°C. Slices were washed in PBS three times (30 min) and then incubated with secondary antibodies for 2 hrs at room temperature. Finally, slices were incubated with Hoechst for 15 min and then stored in PBS at 4 °C. Immunostained slices were observed using a Zeiss LSM-780 inverted confocal laser scanning microscope. Images were analysed with Fiji/ ImageJ.

### Bioinformatic analysis of Human RNA-seq data

The data set was obtained from the open source GEO dataset: GSE97358, which was published through the research of Schafer et al. (Schafer et al., 2017) and GSE133017 which was published through the research of Chen et al. (Chen et al., 2019). This expression data set consists of RNA-sequencing in 168 primary cardiac FB derived from patients (84 control samples and 84 treated with TGF-β). The data set consisted of 64254 genes and was uploaded to R (v4.0.2) and then transformed into expression data using the edgeR package (Robinson et al., 2010). The resulting matrix was filtered to remove minimally-expressed genes. Genes with a log_10_(expression) value less than 0 in at least 90% (>74) of all samples were removed. The resulting dataset contained 14203 genes. The filtered dataset was then separated by culture conditions (control and TGF-β treated). Using the ‘gtools’ package we were able to compare (TGF-β treated vs. control) the expression fold change of all expressed genes in the 84 paired samples (Warnes et al., 2015). The average expression fold change of each gene was then calculated (Log_2_(expression fold change)) and applied to the network described below (all NaN and Inf values were replaced with 0). We then created a correlation matrix of genes which will include; correlation coefficients, p-values and false discovery rate (FDR) using the Hmisc package (Harrell FE and Dupont, 2007). We extracted a list of 14203 genes and their correlation data for the IL11 gene (Supplementary data 1A) and then removed matches with FDR greater than 0.05 (Supplementary data 1B), resulting in 4853 genes with significant adjusted p-values. To further filter our data, we applied another threshold based upon correlation coefficients (where r≥ ±0.5) (Supplementary data 1C), in this dataset values varied between -0.78 and +0.75. The majority of our results were observed to have a low to moderate correlation. For this reason, we applied a cut off for values with a low correlation value. We considered values ≥ ±0.5 to have a strong correlation (Mukaka, 2012). This analysis identified 677 genes which were used to build a protein-protein network. In order to cover other fibrotic related genes we united our IL-11 co-expression network with a recently constructed WWP2 network (683 genes), from cluster of genes which was found to be important in fibrosis and responsible for cell adhesion (Chen et al., 2019).

### Gene set enrichment analysis

In order to reveal the enriched pathways associated with the IL-11 co-expression network, we conducted gene set enrichment analysis using DAVID 6.8 (Huang et al., 2009b; a). Ensemble gene ids were uploaded as a gene list for Homo Sapiens with the same background. For each gene, we searched the database annotations for Gene Ontology, GO_BP_Direct and in Pathways, KEGG_Pathway. We set up the database to only highlight terms with p-value< 0.05 (FDR and fold enrichment were also included added as additional search terms). From this we produced an IL-11 co-expression network (Supplementary data 2).

### Protein-protein interaction network construction

In this study, we aim to construct a protein-protein interaction network based upon genes identified from the IL-11 co-expression network, we also included GPBAR1, which encodes for the bile acid receptor TGR5. To construct this network we used STRING v.11 (‘Search Tool for Retrieval of Interacting Genes/Proteins’) (Szklarczyk et al., 2019) which links proteins based on reported associations between them. In order to establish protein associations, we investigated 7 active interaction sources: textmining, experiments, databases, co-expression, neighbourhood, gene fusion and co-occurrence. Each edge was then ranked based upon the confidence of the protein-protein association being true. During the construction of our protein-protein network, we searched all 7 active sources and only included pairs of genes which showed greater than ‘medium’ confidence behind their association (confidence > 0.4). The resulting network was exported as a table (Supplementary data 3), genes were assigned the average log2 (expression fold change) from filtrated IL-11 dataset.

### Network analysis with Cytoscape 3_7_1

The network which was generated in STRING v.11 was imported in Cytoscape 3_7_1 to generate a network visualization graph (Shannon, 2003) network analysis was performed using NetworkAnalyzer (Assenov et al., 2008). In the resulting network graph (Fig. 6A), node colour represents the average Log_2_(expression fold change) in UDCA vs IL-11 conditions. The size of each node represents the degree of connectivity (i.e. the number connections with other elements of a network). We also identified how GPBAR1 interacts with our combined fibrotic network using the Cytoscape App Pesca3.0.8. This app analyses shortest path from one node (i.e. GPBAR1) to any other element of a network and can be used to determine shortest path between two nodes (Scardoni et al., 2016). We finally classified path lengths as first or second level neighbours of GPBAR1 in order to identify how TGR5 and its downstream signalling can attenuate cardiac fibrosis. In order to examine how IL-11 or UDCA treatment affected our resulting network we did enrichment analysis of the network with WebGestalt. The IL-11 and WWP2/ TGF-β networks were uploaded separately. Following this we tested the networks with gene lists comparing the IL-11 vs. Control and UDCA vs. IL-11 conditions (Fig. 7D and E). Significantly regulated genes were defined as having a false discovery rate <0.05.

### Statistical analysis of experimental data

Data is presented in text as mean ±SEM. Bar charts are presented as mean and SEM of each experimental group. Statistical significance of experiments, unless stated, was determined by one-way ANOVA with Tukey’s post-hoc test. Levels of significance were; *p< 0.05, **p< 0.01, ***p< 0.001. n= number of experiments, N= number of animals/ patients. All experiments were completed in the same laboratory.

Statistical methods used in the bioinformatic analysis of human RNA-seq data are detailed in the methods above.

### Data availability statement

The authors confirm that the data supporting the findings of this study are available within the article and its supplementary materials.

## Supplemental material

Supplementary data 1: IL-11 differential co-expression network construction.

Supplementary data 2: Enrichment analysis for IL-11 and WWP2/ TGF-β differentially co-expressed network.

Supplementary data 3: Protein –protein interaction network for IL-11 and WWP2/ TGF-β co-expressed genes.

Supplementary data 4: Raw data

## Acknowledgments

We wish to thank Mr. Peter O’Gara for his isolation of rat fibroblasts. We thank Kristina Schoonjans for the donation of the TGR5 KO mouse colony. We thank the patients and families for the kind donation of human samples.

This research was funded by Heart Research UK (RG2666/17/19). RC was funded by the BHF project grant PG/16/17/32069.

